# An Early SARS-CoV-2 Omicron Outbreak in a Dormitory in Saint-Petersburg, Russia

**DOI:** 10.1101/2022.11.23.22282648

**Authors:** Galya V. Klink, Daria M. Danilenko, Andrey B. Komissarov, Nikita Yolshin, Olga V. Shneider, Sergey Shcherbak, Elena Nabieva, Nikita Shvyrev, Nadezhda Konovalova, Alyona Zheltukhina, Artem Fadeev, Kseniya Komissarova, Andrey Ksenafontov, Tamila Musaeva, Veronica Eder, Maria Pisareva, Petr Nekrasov, Vladimir Shchur, Georgii A. Bazykin, Dmitry Lioznov

## Abstract

The Omicron variant of SARS-CoV-2 has rapidly spread globally in late 2021 - early 2022, displacing the previously prevalent Delta variant. Before December 16, 2021, community transmission had already been observed in tens of countries globally. However, in Russia, the majority of reported cases at that time had been sporadic and associated with travel. Here, we report an Omicron outbreak at a student dormitory in Saint Petersburg between December 16 - 29, 2021, which was the earliest known instance of large-scale community transmission in Russia. Out of the 465 sampled residents of the dormitory, 180 (38.7%) tested PCR positive. Among the 118 residents for whom the variant has been tested by whole-genome sequencing, 111 (94.1%) carried the Omicron variant. Among these 111 residents, 60 (54.1%) were vaccinated or had reported previous COVID-19. Phylogenetic analysis confirmed that the outbreak was caused by a single introduction of the BA.1.1 sublineage of Omicron. The dormitory-derived clade constituted a significant proportion of BA.1.1 samples in Saint-Petersburg and has spread to other regions of Russia and other countries. The rapid spread of Omicron in a population with preexisting immunity to previous variants underlines its propensity for immune evasion.

## Introduction

The Omicron variant of SARS-CoV-2 was first reported in South Africa on November 24, 2021 (1,2) and has been observed to rapidly spread globally soon thereafter. By mid-December, it outpaced the preceding diversity (mostly Delta) in many countries, including South Africa, the United Kingdom, Australia and Canada, and became the prevalent variant (3). While between May and December 2021, the SARS-CoV-2 epidemic in Russia was dominated by the Delta variant, with one particular Delta lineage, AY.122, having over 90% prevalence (4), by the end of January 2022, Omicron became the dominant variant in Russia as well (https://www.interfax.ru/russia/818539). The details of its onset in Russia are poorly studied. Here, we report an outbreak of Omicron in a student dormitory in the early weeks of the Omicron wave in Russia.

Among the 19 full-genome Omicron samples obtained in Russia and deposited to GISAID by September 1st, 2022 with sampling dates between December 3□15, 12 were from people with known history of travel: 10 to the Republic of South Africa (all sampled on December 3 in Moscow), one to the Dominican Republic (sampled on December 13 in Saint Petersburg), and one to The Republic of the Congo (sampled on December 10 in Rostov-on-Don). Among the seven early genomic samples without known travel history, six were not associated with any other Russian sequences when placed on the UShER phylogenetic tree using on-line UShER tool (5), i.e., represented Russian singletons (6); the seventh sequence was phylogenetically adjacent to the Rostov-on-Don sample with travel history to The Republic of the Congo. Therefore, community transmission of the Omicron variant, if present, was low-level on those dates. Three of the 19 samples belonged to the BA.1.1 lineage, including that from the traveler to The Republic of the Congo.

While the Delta epidemic continued in Saint Petersburg throughout late 2021, with an average of 48.7 daily reported cases per 100K in November (https://xn--80aesfpebagmfblc0a.xn--p1ai/information/), we started systematic screening for early Omicron detection using the Ins214EPE assay (7) on general population samples obtained from multiple hospitals and outpatient clinics. Between November 29 □December 15, we screened 200 to 1000 samples daily.

## Results

On December 16, in the course of screening, we detected Omicron in a hospital sample from a patient without travel history. Follow-up contact tracing revealed that this sample came from a SARS-CoV-2 outbreak in a student dormitory in Saint Petersburg. Between December 17□29, we performed follow-up testing of dormitory residents. Out of the 465 residents, 180 (38.7%) tested positive for COVID-19 over these dates. For 137 samples, the Ins214EPE assay indicated that they were of the Omicron variant.

We performed whole-genome sequencing (WGS) for 118 samples with sufficiently low ct values. 111 of the 118 sequences (94.1%) were classified as Omicron on the basis of WGS. The remaining 7 sequences were classified as non-Omicron (Delta).

### Phylogenetic distribution of samples indicates a single introduction into the dormitory

All seven Delta samples belonged to AY.122, the predominant lineage in Saint Petersburg. Four of them formed a compact clade (transmission lineage (6)), while the remaining three were phylogenetically distinct (singletons (6)). The fact that the Delta samples were scattered across the phylogeny of AY.122 is consistent with multiple distinct sources of non-Omicron infection, in line with a high prevalence of Delta at Saint Petersburg on those dates.

By contrast, all the 111 Omicron samples belonged to the BA.1.1 sublineage, and had a compact phylogenetic distribution within this sublineage (Fig. 1). This is consistent with a single introduction and subsequent spread within the dormitory or multiple infections from a single source.

**Fig. 1.**
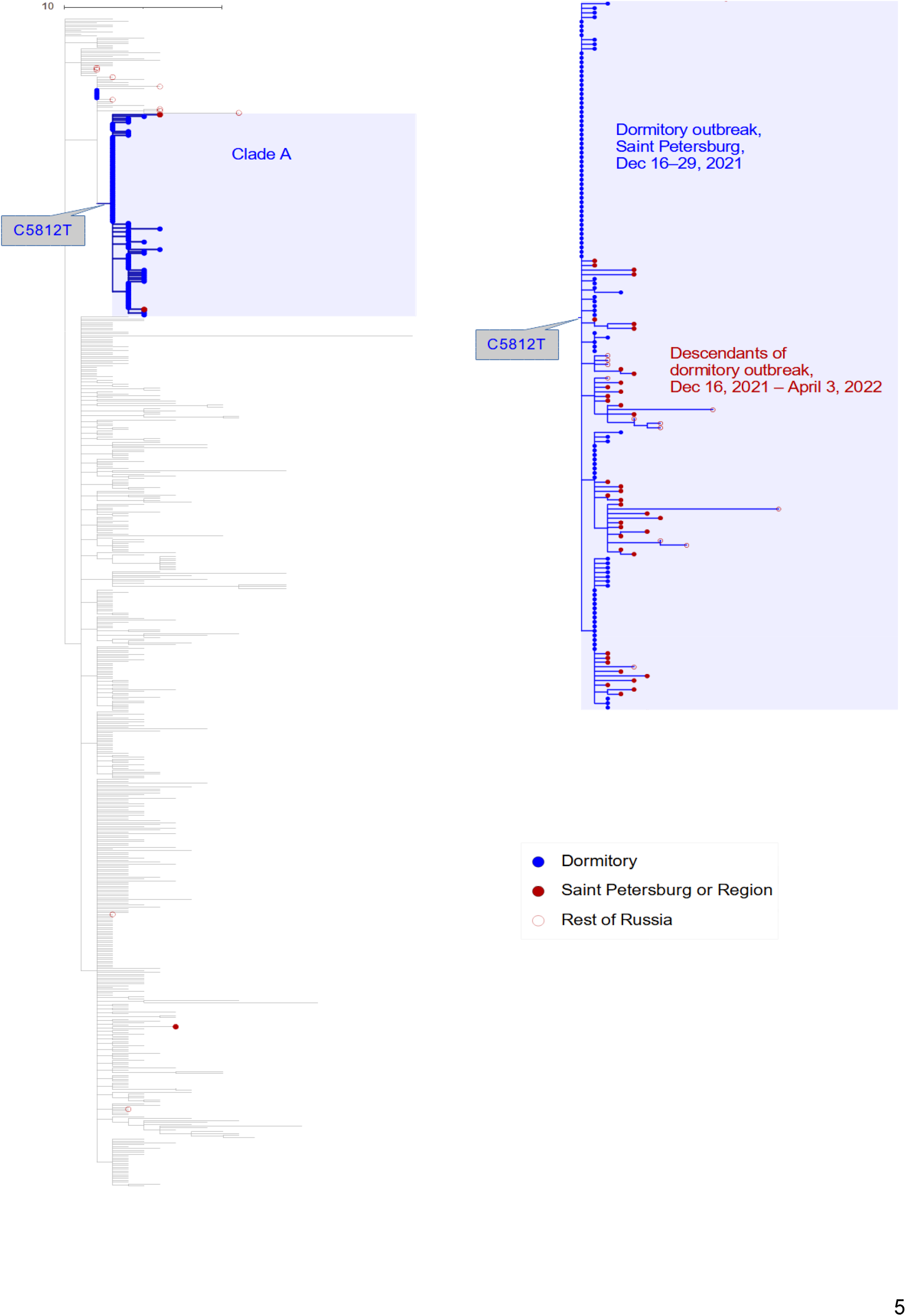
The Russian samples obtained between December 3□30 and the Saint Petersburg dormitory outbreak on the global tree of BA.1.1. All GISAID samples from Russia are shown (in red), together with a random sample of 400 (out of 8396) GISAID Omicron samples obtained in other countries (gray). The clade A defined by the presence of the C5812T mutation is shown in blue. A zoom in of clade A including the outbreak samples (blue) is shown at the right, together with the descendant non-dormitory samples (red).

The BA.1.1 sublineage is characterized by the S:R346K mutation. S:346 is an important immunogenic residue, and various mutations at it allow the virus to escape neutralization by multiple antibodies (8). This site was shown to experience positive selection within the BA.1 sublineage of Omicron (9). However, the arginine-to-lysine change observed in BA.1.1 is chemically conservative, does not lead to a major shift in antibody recognition, and does not confer a significant transmission advantage (10). Therefore, the fact that the dormitory outbreak has been caused by BA.1.1 rather than the ancestral BA.1 lineage is probably due to a founder effect. In any case, the extensive spread of a single Omicron sublineage but none of the three Delta sublineages is consistent with a higher transmission rate of Omicron compared to Delta in this setting.

All but five dormitory samples formed a single compact clade within BA.1.1, which we refer to as clade A. This clade was characterized by the C5812T synonymous mutation (nsp3:D1031D). Notably, the remaining five dormitory samples were positioned at the root of clade A, i.e., carried all mutations of clade A except C5812T; however, even for these samples, position 5812 was polymorphic, with derived variant T present at between 7-50% of sequencing reads, suggesting that the C5812T mutation arose in the dormitory at the beginning of the outbreak.

We characterized the introduction and transmission of the virus in the dormitory outbreak using a phylodynamics approach. For this, we applied the birth-death skyline model (11) of BEAST2 (12) to the dormitory samples of Omicron. We considered three different fixed values of clock rate: 0.75x10^−3^, 0.95x10^−3^ and 1.15x10^−3^ (1,13). In each scenario, the effective reproductive number R_e_ was estimated for three time periods: R_1_ before the first sample was collected on December 16, R_2_ between December 16 and December 24, and R_3_ between December 24 and December 29 (the date of the last sample collected).

The most recent common ancestor of the dormitory outbreak is estimated to be on December 2 with the 95% CI [Nov 23, Dec 9] for the lowest value of clock rate (0.75x10^−3^), December 5 [Nov 28, Dec 11] for the intermediate value (0.95x10^−3^), and December 7 [Dec 1, Dec 12] for the highest value of 1.15x10^−3^ (Fig. 2, Supplementary Table 1). Assuming a single introduction into the dormitory which is supported by the monophyly of the dormitory samples, this implies that the infection was introduced into the dormitory about two weeks prior to the collection of the first sample on December 16. The initial effective reproductive number R_1_ is high in all three scenarios: 3.90 with 95% CI of [2.22, 5.67], 4.59 [2.56, 6.96], or 5.23 [2.82, 8.00] for different clock rate values respectively. Later, it drops by a factor of approximately 2.5 consistently in all runs, with R_2_ being equal to 1.69 [1.00, 2.41], 1.83 [1.11, 2.58] and 1.97 [1.21, 2.74] respectively. R_3_ has a very wide credible interval which includes R_e_=1 and it is not informative about the phylodynamics between December 25 and December 29. This is explainable by the low number of samples from this time period.

**Fig. 2.**
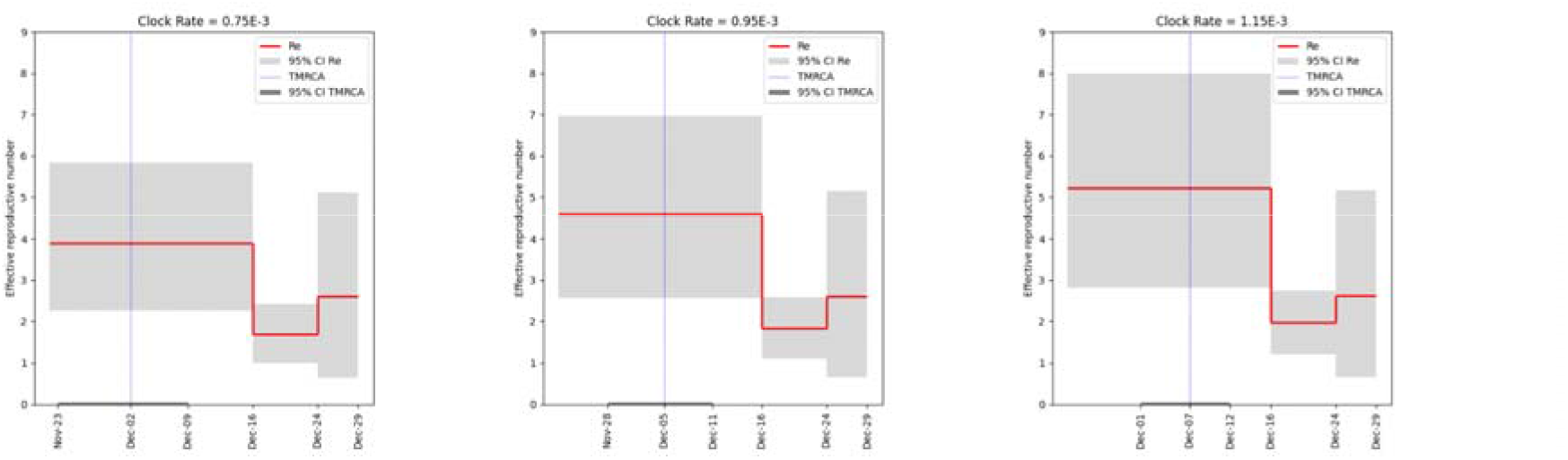
Skyline plots for the effective reproductive number R_e_ for different values of molecular clock rate.

**Supplementary Table 1**. Phylodynamic parameters for the dormitory outbreak inferred for three values of clock rate under birth-death skyline model.

The substantial fraction of infected individuals in the dormitory outbreak likely had some preexisting immunity to SARS-CoV-2. Among the 137 patients who tested positive for Omicron, 71 (51.8%) reported previous infection or vaccination. This is in line with the high immune evasion properties of Omicron (14,15).

### An elevated rate of within-room transmission

The dormitory occupied a single multistory building. Most dormitory rooms had a 4-bed layout, with up to 24 rooms per floor. For 104 of the 111 Omicron-positive residents, the floor and room were known. We asked how the risk of transmission was affected by living together in the same room or on the same floor with an infected individual. We reasoned that if the Omicron variant has been introduced into the dormitory just once, all differences between samples originated during within-dormitory transmission. Therefore, the samples separated by a direct transmission of the virus will be distinguished by fewer differences, compared to samples separated by a chain of more than one transmission through other individuals.

To test this, we calculated the mean pairwise phylogenetic distance *m* (which typically equaled the number of single-nucleotide differences) between samples from individuals residing in the same room or at the same floor, and compared it with the expected distance between samples from the same floor or from anywhere in the building. To obtain these expected values, we reshuffled across individuals the room labels while controlling or not controlling for the floor; or the floor labels.

We found that when two infected individuals resided in the same room, the phylogenetic distance between their SARS-CoV-2 samples was 1.8 times lower compared to an average pair of infected individuals residing on any floor (0.65 vs. 1.19, Fig. 3A), and 1.5 times lower compared to individuals residing on the same floor (0.65 vs. 0.99, Fig. 3B), and these differences were significant (p=0.0001 and p=0.006 respectively). Conversely, accommodation on the same floor irrespective of the room did not lower the phylogenetic distance between samples, compared to pairs of infected individuals from anywhere in the building (1.18 vs 1.18, p=0.491, Fig. 3C). These results indicate that residing in the same room with an infected individual increased the risk of transmission from that individual, while living on the same floor but in a different room had no effect.

**Fig. 3.**
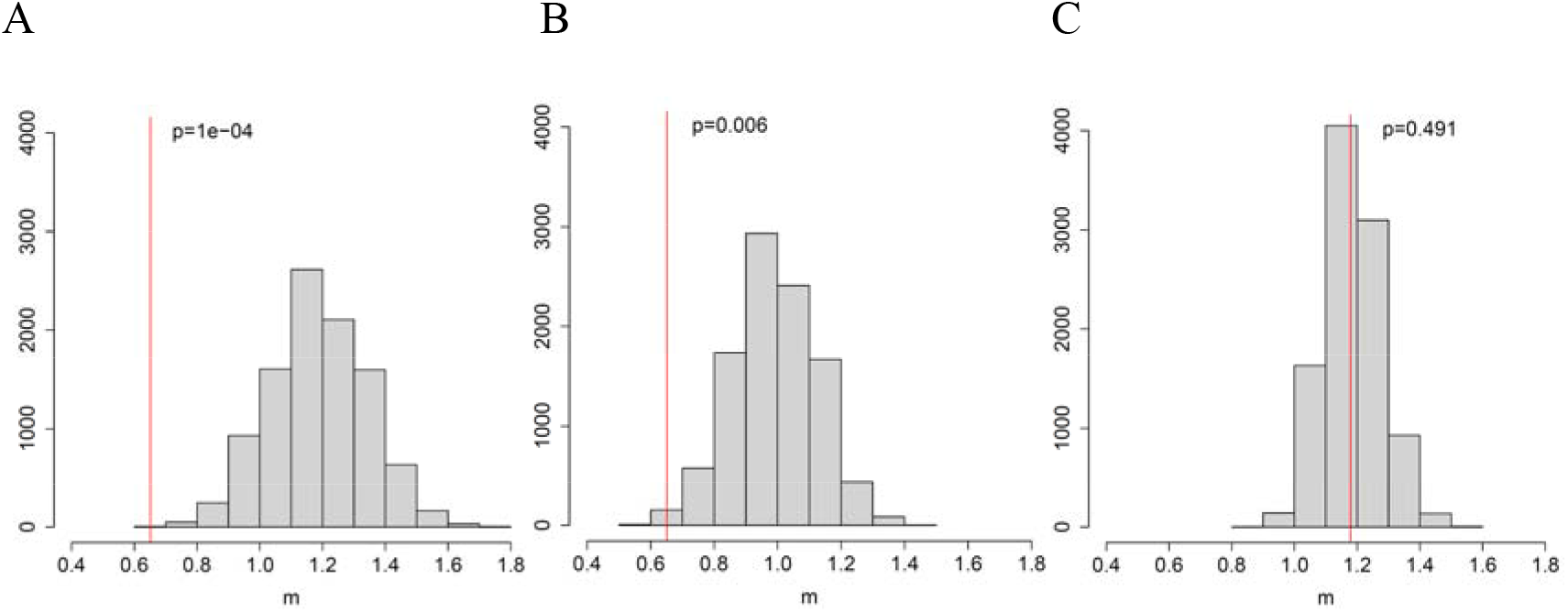
The mean phylogenetic distance *m* between two samples from the same room (A, B) or floor (C) (red), compared to the expected distributions obtained by reshuffling of room labels independent of the floor (A), within the floor (B), or by reshuffling of floor labels (C) of samples. p, fraction of reshuffling trials with mean phylogenetic distance below *m*.

### The role of dormitory outbreak in the Russian and global epidemic of Omicron

The BA.1.1 lineage comprised a considerable fraction of Russian samples in the beginning of 2022 (Fig. 4A). The UShER tree of BA.1.1 contained 489 non-dormitory Russian samples obtained after December 16, 2021. Among them, 51 (10%; Wilson 95% CI = 8%-13%) belonged to clade A and carried all three of its characteristic mutations (Fig. 1, Supplementary Table 2).

**Fig. 4.**
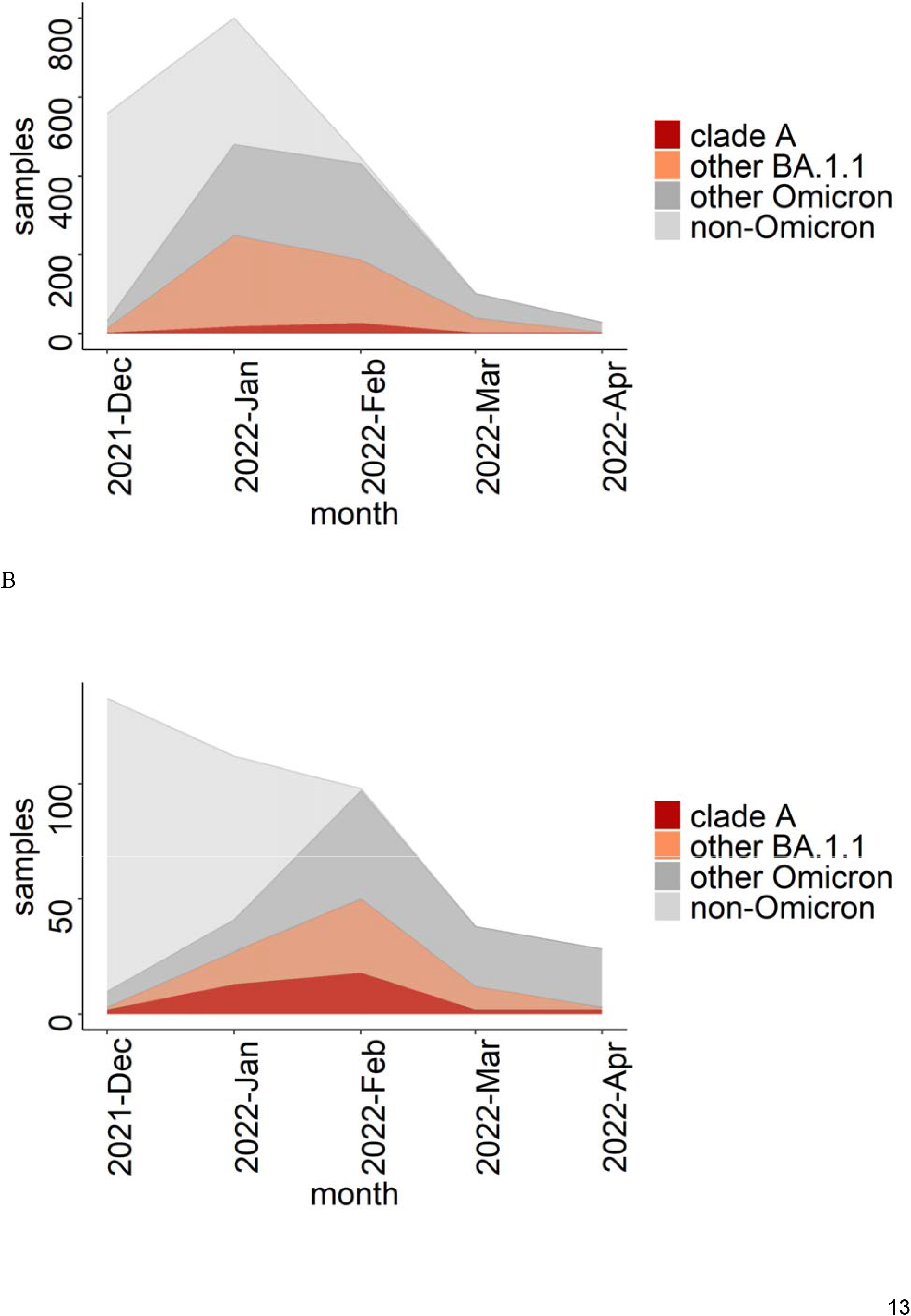
The fraction of clade A, BA.1.1 and Omicron samples among Russian (A) and Saint-Petersburg (B) samples from GISAID included in the UShER phylogenetic tree downloaded on May 26, 2022. Samples from the dormitory are not included.

Among the dormitory samples of clade A, 47.2% (50/106) of which were basal, i.e. carried no extra changes on top of the characteristic C5812T mutation of clade A. By contrast, all 51 non-dormitory samples carried extra mutations, indicating that they were exported from the dormitory into the general population of Saint Petersburg and beyond. According to the phylogenetic tree, there were at least three such exports of clade A (Fig. 1). Clade A samples were most frequent in Saint Petersburg (comprising 18.6% of all Omicron samples in February 2022) as well as the surrounding Leningrad Oblast (Figs 4B and 5), pointing to a considerable contribution of the dormitory outbreak to the Omicron wave here. Meanwhile, the role of the dormitory outbreak in the spread of Omicron in most other Russia’s regions was negligible: for example, none of the 81 samples from Moscow belonged to clade A (Fig. 5).

**Fig. 5.**
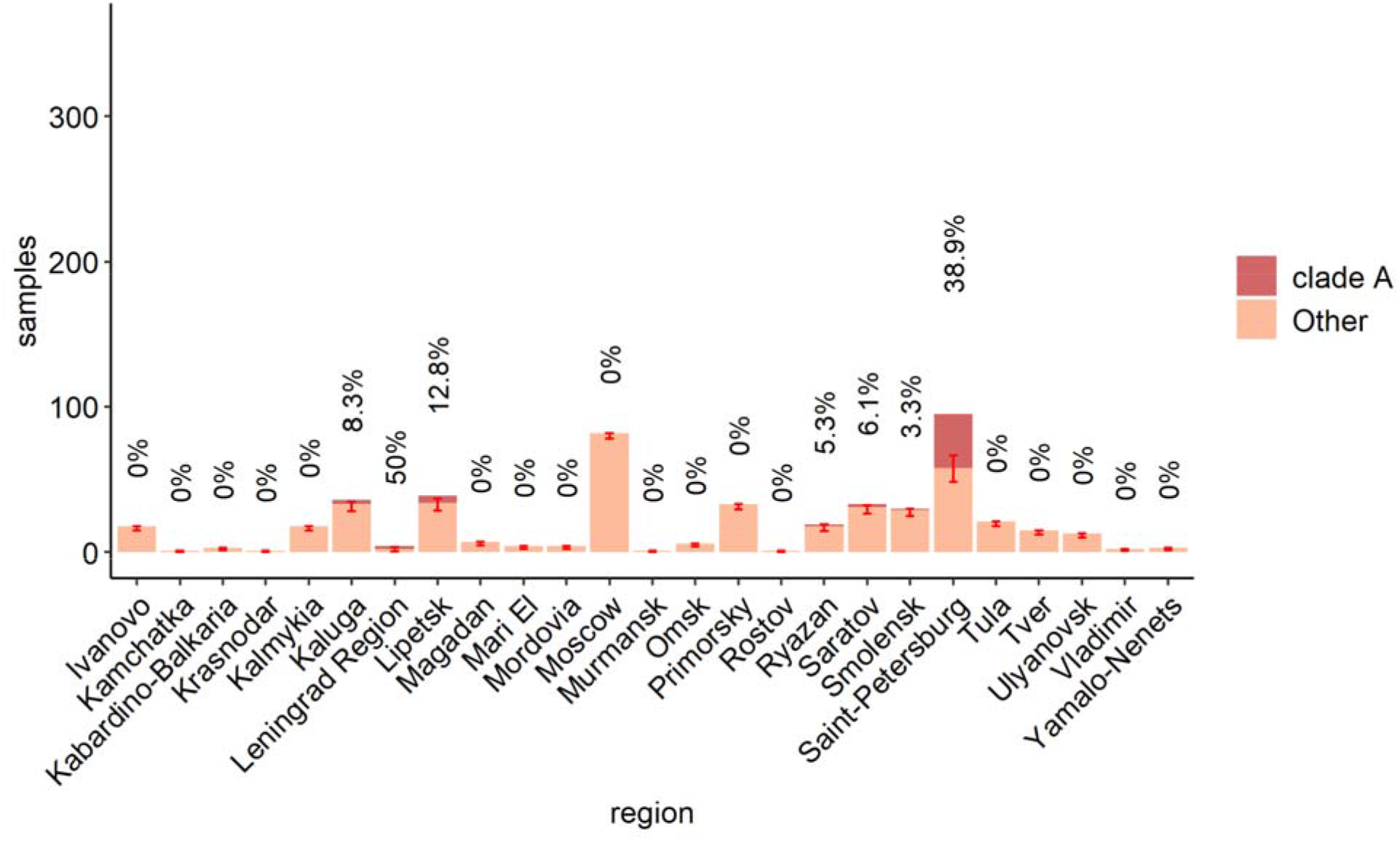
Clade A samples (orange) among all BA.1.1 samples in Russia’s regions. Numbers on bars are the percentage of clade A samples in each region; 95% Wilson CIs are shown as bars. Dormitory samples are not included.

**Supplementary Table 2**. The fraction of clade A samples among all Russian BA.1.1 samples obtained after 16th December 2021 that were present in the UShER tree downloaded on May 25, 2022. *95% Wilson CI

Besides samples from Russia, clade A also carried 118 non-Russian samples from 20 countries (Supplementary Table 3). All of them were collected after December 16, 2021. The fraction of such samples among all BA.1.1 samples was low (<<1%) in all countries except Estonia where it reached 1.8%. Notably, the two countries with the highest fraction, Estonia (1.8%; Wilson 95% CI = 0.8%-3.9%) and Finland (0.5%; Wilson 95% CI = 0.3%-0.8%), are geographically close to Saint Petersburg and are frequent travel destinations for Saint Petersburg residents.

**Supplementary Table 3**. The fraction of clade A samples among all non-russian BA.1.1 samples obtained after December 16, 2021. Only countries with non-zero fraction are shown.*95% Wilson CI

## Discussion

In this work, we describe an outbreak of BA.1.1 in a student dormitory in Saint Petersburg at the beginning of the wave caused by Omicron. We show that the dormitory-derived variant spilled over into the general population of Saint Petersburg, representing a substantial fraction among the BA.1.1 samples here. Additionally, it spread to some other regions of Russia and to other countries. As clade A differs from the root of BA.1.1 in three nucleotide mutations, all of which are synonymous, it is unlikely that it had a fitness difference, meaning that most likely it has spread due to chance. Transmission of SARS-CoV2 is highly non-uniform, providing an important role for superspreading events in the epidemic (16,17). Recently, using early Omicron transmission chains in Hong Kong, it was estimated that 80% of transmissions were generated by 20% of cases, and the superspreading potential of Omicron was suggested to be higher than for the variants circulating in 2020 (18). In Russia, the whole Delta wave was mostly made by a single clade that has likely spread due to chance (4).

Hotels and dormitories provide a major potential for superspreading. For Omicron, it was shown that even in a quarantine hotel, the virus moved between neighboring rooms by circulating air (19). Nevertheless, in this study, we show that even in a student dormitory, where residents of different rooms are likely to actively communicate with each other, infection from roommates were more likely than from other residents of the same floor or the entire building. Therefore, living places with layouts carrying more beds per room may host more rapidly growing outbreaks than places with smaller rooms.

At the onset of Omicron in South Africa, its estimated per day growth advantage over Delta was estimated to be 0.24 (1). In agreement with this, while we detected four independent introductions of Delta in the dormitory simultaneously with the introduction of Omicron, neither of these four introductions led to an outbreak. The higher transmissibility of Omicron is thought to be mainly due to its immune evasion properties (14,15). In our study, near half of dormitory residents with Omicron variant were previously vaccinated or infected, illustrating its potential for spread in a population with preexisting partial immunity.

Notably, despite the high fraction of dormitory-derived clade A in Saint Petersburg and Leningrad Oblast, its prevalence in other regions of Russia was low (Fig. 5), pointing out that even close regions with a high passenger flow between them, such as Saint Petersburg and Moscow, have unique epidemiological histories of equally fit viral variants. This is of course conditional on similar fitness of these variants; a novel advantageous variant can rapidly spread across a country and the world, as repeatedly observed during the SARS-CoV-2 pandemic.

## Methods

We performed whole-genome sequencing (WGS) using the SARS-CoV-2 ARTIC V4 protocol and the Oxford Nanopore gridION or Illumina NextSeq 2000 sequencing technology. Consensus genome assembly was performed by bwa-mem and bcftools, preceded by adapter and primer trimming by trimmomatic, ivar (Illumina) or BAMClipper (Oxford Nanopore) and custom scripts. An alternative allele was called if its read frequency exceeded 0.5 at a position. Positions with coverage below 10 (Illumina) or 20 (Oxford Nanopore) were masked as N.

A few dormitory sequences assigned to the Omicron lineage had positions that were called as ancestral and/or Delta nucleotides. While these could be legitimate new mutations (including reversions), a close analysis of the NGS data hinted at the possibility that these could be artifacts of primer integration into reads (leading to the reference variant) or contamination (or coinfection) with the Delta variant. To be on the conservative side, we marked such positions as N in the dormitory sequences for the purposes of tree construction.

For phylogenetic analysis, we downloaded the UShER SARS-CoV2 phylogenetic tree on May 26th, 2022 and extracted a subtree of 572,763 BA.1.1 samples available in GISAID. We removed all dormitory samples from the tree and added their improved consensus to this tree with the UShER tool (5) and visualized it with iTOL (20). The same was done with seven Russian samples of Delta and Delta subtree. Statistical analysis was performed with R (21). Wilson confidence intervals were calculated with Hmisc package (22), and plots were made using tidyverse (23) and ggsignif (24) packages for R.

## Supporting information

GISAID acknowledgements

## Data Availability

All data produced in the present work are contained in the manuscript

## Acknowledgements

We thank UShER team for providing currently updating phylogenetic tree of SARS-CoV2. We are grateful to all GISAID submitting and originating labs (Supplementary File 1) for rapid open release of SARS-CoV-2 sequencing data.

Samples acquisition and sequencing were supported by the Ministry of Health of Russian Federation as a research work of the state assignment #121110800170-8 (2022). Bioinformatic analysis was funded by the Russian Science Foundation (project no. 21-74-20160 to G.A.B.)

## Notes

### Competing Interest Statement

The authors have declared no competing interest.

### Author Declarations

Samples used in this study were collected as part of approved ongoing surveillance conducted by the Smorodintsev Research Institute of Influenza. Written informed consent was obtained from all subjects. All samples were deidentified prior to receipt by the study team. The study was presented to the Local Ethics Committee at the Smorodintsev Research Institute of Influenza. The Committee concluded (protocol #177) that the study does not make use of new identifiable biological samples and does not bring forward any new sensitive data. Therefore, according to the rules of the Committee and national regulations this project does not require ethical approval.

