## Supplementary Table1-3 for "An Early SARS-CoV-2 Omicron Outbreak in a Dormitory in Saint-Petersburg, Russia"

**Supplementary Table 1.** Phylodynamic parameters for the dormitory outbreak inferred for three values of clock rate under birth-death skyline model.

| Clock  rate | 7.25E-4  [6.29E-4, 8.19E-4] | 9.16E-4  [8.20E-4, 1.01E-3] | 11.1E-4  [10.16E-4, 12.09E-4] |
| --- | --- | --- | --- |
| Likelihood | -40833.24 | -40835.25 | -40836.99 |
| Prior | 782.24 | 798.83 | 811.07 |
| TMRCA | 1 December  [22 November,  9 December] | 4 December  [26 November,  11 December] | 6 December  [1 December  11 December] |
| Re1 | 3.79  [2.22, 5.67] | 4.47  [2.48, 6.85] | 5.08  [2.77, 7.86] |
| Re2 | 1.67  [0.99, 2.39] | 1.80  [1.10, 2.55] | 1.94  [1.20, 2.72] |
| Re3 | 2.61  [0.66, 5.18] | 2.61  [0.65, 5.17] | 2.61  [0.64, 5.14] |
| Sp1 | 0 | 0 | 0 |
| Sp2 | 0.90  [0.71, 1] | 0.91  [0.75, 1] | 0.92  [0.78, 1] |
| Sp3 | 0 | 0 | 0 |
| Sp4 | 0.27  [0.02, 0.60] | 0.27  [0.01, 0.58] | 0.26  [0.01, 0.58] |

**Supplementary Table 2.** Fraction of descendants of dormitory clade among all Russian BA.1.1 samples obtained after 16th December 2021 that were present on the phylogenetic tree provided by UShER on 25th May 2022. *95% Wilson CI

| Region | All  Samples | Dorm-derived  Samples | Dorm-derived  Fraction | Lower  Bound | Upper  Bound |
| --- | --- | --- | --- | --- | --- |
| LEN | 4 | 2 | 0.5 | 0.15 | 0.85 |
| SPE | 100 | 41 | 0.41 | 0.32 | 0.51 |
| LIP | 39 | 5 | 0.13 | 0.06 | 0.27 |
| KLU | 36 | 3 | 0.08 | 0.03 | 0.22 |
| SAR | 33 | 2 | 0.06 | 0.02 | 0.2 |
| RYA | 19 | 1 | 0.05 | 0 | 0.25 |
| SMO | 30 | 1 | 0.03 | 0 | 0.17 |
| IVA | 18 | 0 | 0 | 0 | 0.18 |
| TUL | 21 | 0 | 0 | 0 | 0.15 |
| YAN | 3 | 0 | 0 | 0 | 0.56 |
| MOW | 81 | 0 | 0 | 0 | 0.05 |
| MO | 4 | 0 | 0 | 0 | 0.49 |
| PRI | 33 | 0 | 0 | 0 | 0.1 |
| ME | 4 | 0 | 0 | 0 | 0.49 |
| KL | 18 | 0 | 0 | 0 | 0.18 |
| TVE | 15 | 0 | 0 | 0 | 0.2 |
| MAG | 7 | 0 | 0 | 0 | 0.35 |
| VLA | 2 | 0 | 0 | 0 | 0.66 |
| KAM | 1 | 0 | 0 | 0 | 0.95 |
| ULY | 13 | 0 | 0 | 0 | 0.23 |
| KB | 3 | 0 | 0 | 0 | 0.56 |
| OMS | 6 | 0 | 0 | 0 | 0.39 |
| KDA | 1 | 0 | 0 | 0 | 0.95 |
| MUR | 1 | 0 | 0 | 0 | 0.95 |

**Supplementary Table 3.** Fraction of descendants of dormitory clade among all non-russian BA.1.1 samples obtained after 16th December 2021. Only countries with non-zero fraction are shown.*95% Wilson CI

| Country | All  Samples | Dorm-derived  Samples | Dorm-derived  Fraction | Lower  Bound | Upper  Bound |
| --- | --- | --- | --- | --- | --- |
| Estonia | 328 | 6 | 0.018 | 0.01 | 0.04 |
| Finland | 2340 | 11 | 0.005 | 0 | 0.01 |
| Austria | 1358 | 2 | 0.001 | 0 | 0.01 |
| Israel | 7087 | 10 | 0.001 | 0 | 0 |
| Argentina | 729 | 1 | 0.001 | 0 | 0.01 |
| Denmark | 2964 | 4 | 0.001 | 0 | 0 |
| Germany | 37666 | 40 | 0.001 | 0 | 0 |
| France | 10414 | 8 | 0.0008 | 0 | 0 |
| Slovakia | 2760 | 2 | 0.0007 | 0 | 0 |
| Norway | 1390 | 1 | 0.0007 | 0 | 0 |
| India | 1517 | 1 | 0.0007 | 0 | 0 |
| Switzerland | 4900 | 2 | 0.0004 | 0 | 0 |
| Slovenia | 6263 | 2 | 0.0003 | 0 | 0 |
| Italy | 3170 | 1 | 0.0003 | 0 | 0 |
| Netherlands | 4448 | 1 | 0.0002 | 0 | 0 |
| Japan | 9015 | 2 | 0.0002 | 0 | 0 |
| Belgium | 4707 | 1 | 0.0002 | 0 | 0 |
| England | 96314 | 18 | 0.0002 | 0 | 0 |
| Scotland | 21704 | 4 | 0.0002 | 0 | 0 |
| USA | 271041 | 1 | 0.000007 | 0 | 0 |
